# Using different epidemiological models to modeling the epidemic dynamics in Brazil

**DOI:** 10.1101/2020.04.29.20085100

**Authors:** Lucas Ravellys Pyrrho de Alcantara, Anderson Rodrigues de Almeida, Maira Galdino da Rocha Pitta, Lucio Camara e Silva, Artur Paiva Coutinho

## Abstract

In this paper we provide forecasts of the cumulative number of confirmed reported cases in Brazil, specifically in Pernambuco and Ceara, by using the generalized logistic growth model, the Richards growth model and Susceptible, Un-quanrantined infected, Quarantined infected, Confirmed infected (SUQC) phenomenological model. We rely on the Nash-Sutcliffe efficiency (NSE), root-mean-square error (RMSE) and mean absolute relative error (MARE) to quantify the quality of the models’ fits during the calibrationAll of these analyzes have been valid until the present date, April 14, 2020. The different models provide insights of our scenario predictions.

## 1. Introduction

According to (Ferguson et al., 2020), the COVID-19 pandemic is now a major global health threat. In Brazil, due concern for the rapid spread of the virus throughout the country, considering human transmission, the Federal and State Governments have adopted containment strategies throughout the whole country. As of 16^th^ April 2020, there have been 28,320 cases, in which 14,026 recovery and 1,736 deaths confirmed in Brazil.

(Liu et al., 2020) stated that several studies have estimated basic production number of novel COVID-19. Mathematical modelling of disease transmission have a greater role in supporting clinical diagnosis and optimizing a combination of strategies (Jia & Lu, 2020) understanding, e.g., i) how transmissible the disease is, ii) when the infectiousness is highest during the course of infection, iii) how severe the infection is, and iv) how effective interventions have been and ought to be (Tang et al., 2020). However, time-varying transmission dynamics of COVID19 during the outbreak remain unclear (Liu et al., 2020).

Most of previous work use simple exponential growth models and focus on the early growing process. On the other hand, there are also many works arguing that the number of infected people follows a trajectory different from a simple exponential growth.

Thus, the present paper aims to use phenomenological models to dissect the development of the epidemics in Brazil, making a forecast of the daily incidence for the next five days and based on a 365-day simulation, starting on March 18, showing the likely size of the outbreak. For this, Richards growth model (Wu, Darcet, Wang, & Sornette, 2020) was used to characterize the dynamics of COVID-19 in Brail, specifically in the state of Pernambuco and Ceara, northeast Brazil. Thus, this paper is an exploratory evaluation in analysis that contribute not only to the literature, but to local governments.

## 2. Methods

### 2.1 Data Collection

As in (Roosa et al., 2020), we obtained daily updates (until April 16, 2020) of the cumulative number of reported confirmed cases for the 2019-nCoV epidemic across states in Brazil, from the Brazilian Ministry of Nacional Health website (saude.gov.br). The data contains information about the 26 states and a Federal district of Brazil. Besides these data, we also collect the measures that each state is taking to combat covid-19 (www.agenciabrasil.ebc.com.br).

### 2.2 Models

At present, several researchers have contributed to the literature about the COVID-19, thus different methodologies have been implemented to support and control the increasing number of infected human (Roda, Varughese, Han, & Li, 2020), (S. Zhang et al., 2020), (Roosa et al., 2020). In this context, there are traditional models, as: SIR (susceptible population (S), Infected population (I), and recovered population (R, Including death) model has been widely used for modeling infectious diseases (Yu, 2020). SEIR (susceptible-exposed-infected-removed) model (Zhou et al., 2020), in which is considered the Exposed, ones.

In this paper, we generate short-term forecasts in real-time using three phenomenological models that have been previously used to derive short-term forecasts: (i) The generalized logistic growth model (GLM) extends the simple logistic growth model to accommodate sub-exponential growth dynamics with a scaling of growth parameter, p. Logistic model (GLM) based on the initial growth phase of an epidemic tend to under predict disease incidence before the inflection point has occurred

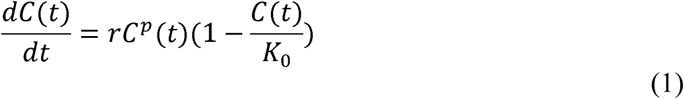

This model also includes flexible growth scaling via the parameter to model a range of early epidemic growth profiles ranging from constant incidence (p=0), polynomial (0 < p < 1) and exponential growth dynamics (p =1). The remaining model parameters are as follows: r is the growth rate, and K_0_ is the final epidemic size. For this model, we estimate Θ= (r, p, K_0_) where f (t, Θ) = C’(t) and fix the initial number of cases C(0) according to the first observation in the data.

(ii) (Wu et al., 2020) generate short-term forecasts in real-time using the generalized Richards model to the reported number of infected cases. The original Richards growth model describes three free parameters, which has been fitted to a range of logistic-type epidemic curves, according to differential Eq. (1) (Chowell, 2017).

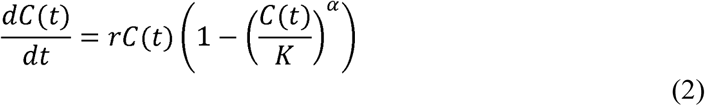

where, where C(t) represents the cumulative number of cases at time t, r is the growth rate in the initial stage and K is the final size of the epidemic. However, according to these authors, during the early stages of disease propagation, when C(t) is significantly smaller than K, this model assumes an initial exponential growth phase. To account for initial sub-exponential growth dynamics, it is incorporated a deceleration of growth parameter (p), *p* ∊ [0.1], allowing the model to capture different growth profiles, replacing the growth term rC by rC^p^. Besides, the α exponent measures the deviation of the s-shaped symmetric dynamics of the simple logistic curve. Basically, the Generalized Richards model (GRM) is defined by the differential Eq. (2):

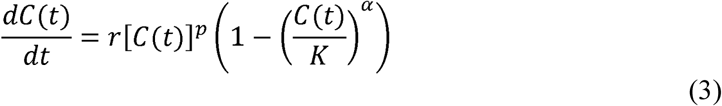

For more detail of this model, we suggest (Wu et al., 2020);(Roosa et al., 2020) (Zhao & Chen, 2020) (Chowell, 2017)

(iii) (Zhao & Chen, 2020) develop a Susceptible, Un-quanrantined infected, Quarantined infected, Confirmed infected (SUQC) model to characterize the dynamics of COVID-19. In this model, S= S(t) is the number of susceptible individuals with no resistance to disease in the population; U=U(t), is the number of infected and un-quarantined individuals that can be either presymptomatic or symptomatic; Q=Q(t), is the number of quarantined infected individuals. C=C(t), the number of confirmed infected cases. Thus, we can developed a composite variable I(t) = U(t) + Q(t) + C(t), representing the real cumulative number of infected individuals at time t. The model comprises the following independent parameters: *α* is the infection rate, the mean number of new infected caused by an un-quarantined infected per day (*α* ∊ [0, *∞*)); *γ*_1_is the quarantine rate for an un-quarantined infected being quarantined, with the range *γ*_1_ ∊ [0,1]; *γ*_2_, the confirmation rate of Q, is the probability that the quarantined infected are identified to be confirmatory cases by a conventional method, such as the laboratory diagnosis, with the range *γ*_2_ ∊ [0,1]. Thus, the ODE equations to model the dynamics of infectious disease and the control by artificial factors (Eq.3)

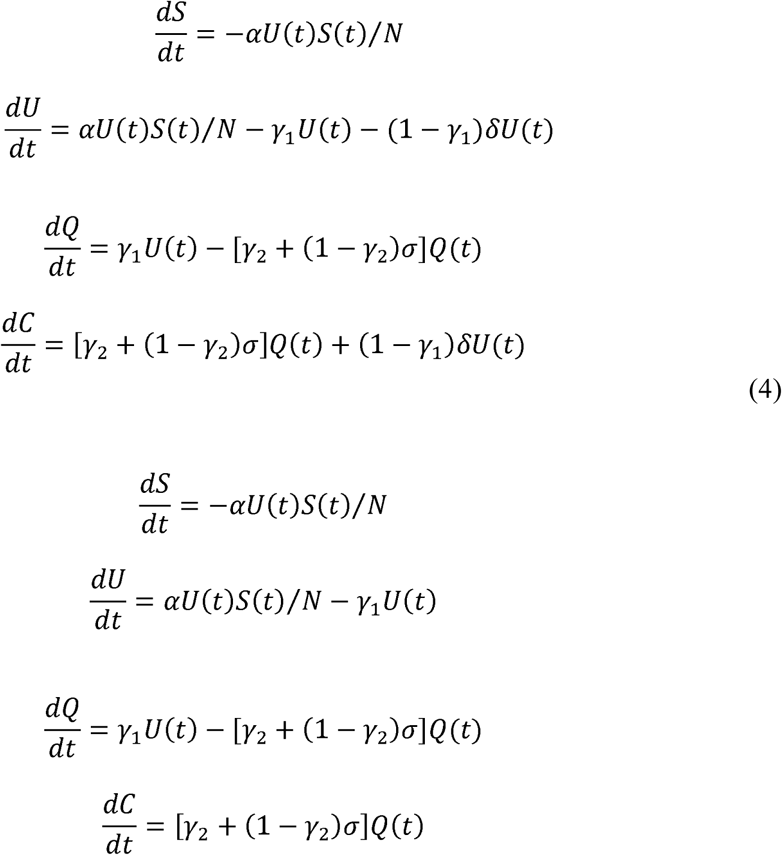

According to the authors, from the above SUQC model, we can further define some biologically meaningful parameters, for monitoring and predicting the trend of disease: T = 1/*γ*1 is the mean waiting time from quarantine to confirmation, w = 1/[*γ*_2_ − (1 − *γ*_2_)*σ*] is the meantime delay from isolation to confirmation; the reproductive number of the infection is R=α/γ1. Differently from SIR/SEIR models, the number of removed individuals, is not included in the model as in the. Once the infected are quarantined, the authors assume their probability of infecting susceptible individuals is zero, and thus no matter the infected are recovered or not, they have no effect on the dynamics of the epidemic system. Besides, with the results of the model, the authors also analyzed the intervention effects of control measures

Therefore, once the SUQC model (Zhao & Chen, 2020) takes into account all the particularities of COVID-19: (i) the epidemic has a probability of infection during the incubation period (pre-symptomatic); (ii) various isolation measures are used to control the development of the epidemic; (iii) the main source of data is the daily number of confirmed infections reported in the official report, affected by the detection method and with a delay between the actual infected number and the confirmed infected number, we agreed that their model is more suitable for analysis than other existing epidemic models, and finally we decided to use it. For more detail of this model, we suggest (Zhao & Chen, 2020)

## 3. Results

### 3.1 Model calibration

We calibrate each model to the daily cumulative reported case counts for Brazil and other states (Pernambuco and Ceara). After fitting the model with the recent data from Brazilian Ministry of Nacional Health website (saude.gov.br), we make a series of predictions about future dynamics of the COVID-19 outbreak in Brazil and the state of Pernambuco. We also extended some analysis for the state of Ceara.

We estimate the best-fit model solution to the reported data using nonlinear least squares fitting. This process yields the set of model parameters Θ that minimizes the sum of squared errors between the model f(t, Θ) and the data y_t_; where Θ_GLM_=(r, p, K), Θ_Rich_=(r, p, K, α), and Θ_SUQC_=(α,β,γ1,S(0),U(0),Q(0),C(0)) correspond to the estimated parameter sets for the GLM, the Richards model, and the SUQC model, respectively.

Table 1, below, shows the GLM model’s parameters.

**Table 1.** Parameter estimation of the epidemic dynamics in Brazil, Ceara, Pernambuco.

|  | **r** | **p** | **K** | **NSE** | **RMSE** | **MARE** |
| --- | --- | --- | --- | --- | --- | --- |
| Brazil | 0.432 | 0.842 | 32433900.646 | 0.991 | 868.387 | 0.027 |
| CE | 1.047 | 0.679 | 1183986.722 | 0.989 | 84.565 | 0.037 |
| PE | 1.361 | 0.695 | 1524686.862 | 0.995 | 57.662 | 0.039 |

Table 2, below, shows the Richards model’s parameters.

**Table 2.** Parameter estimation of the epidemic dynamics in Brazil, Ceara, Pernambuco.

|  | **r** | **p** | **K** | **$\alpha$** | **NSE** | **RMSE** | **MARE** |
| --- | --- | --- | --- | --- | --- | --- | --- |
| Brazil | 0.527 | 0.855 | 32433901.827 | 0.171 | 0.993 | 774.540 | 0.025 |
| CE | 1.746 | 1.000 | 1183991.236 | 0.009 | 0.990 | 81.428 | 0.033 |
| PE | 0.997 | 0.764 | 1524337.774 | 0.265 | 0.993 | 67.969 | 0.046 |

Table 3, below, shows the SUQC model’s parameters.

**Table 3.** Parameter estimation of the epidemic dynamics in Brazil, Ceara, Pernambuco.

|  | **r** | **$\alpha$** | **$\beta$** | **$\gamma_1$** | **$S_{(0)}$** | **$U_{(0)}$** | **$Q_{(0)}$** | **$C_{(0)}$** | **NSE** | **RMSE** | **MARE** |
| --- | --- | --- | --- | --- | --- | --- | --- | --- | --- | --- | --- |
| Brazil | 4.050 | 0.212 | 0.145 | 0.052 | 119586642.484 | 37466.894 | 6843.530 | 11130.000 | 0.982 | 1241.051 | 0.037 |
| CE | 4.961 | 0.251 | 0.125 | 0.051 | 5127326.325 | 2635.755 | 848.377 | 823.000 | 0.992 | 72.440 | 0.030 |
| PE | 1.892 | 0.308 | 0.223 | 0.163 | 6900065.105 | 911.134 | 204.208 | 201.000 | 0.999 | 26.163 | 0.018 |

It is noted that the state of Pernambuco had the lowest detection rate of the virus, this was possible due to the lack of a virus detection test in the state. However, with the arrival of the new tests, it is expected that this rate will increase.

### 3.2 Initial Growth Rate (r) and Final magnitude of the epidemic (K) Analysis

In Figures 1–2 the growth rate of confirmed cases in whole Brazil are presented, based on Generalized Richards model and with the standard logistic growth model. While the two models estimated stable and nearly equivalent growth rates in Brazil, the estimated growth rates for other states vary across models and do not follow a distinct trend as more data become available. However, the scaling and size parameters (Figure 3–4) remain relatively stable across all states.

**Fig 1.**
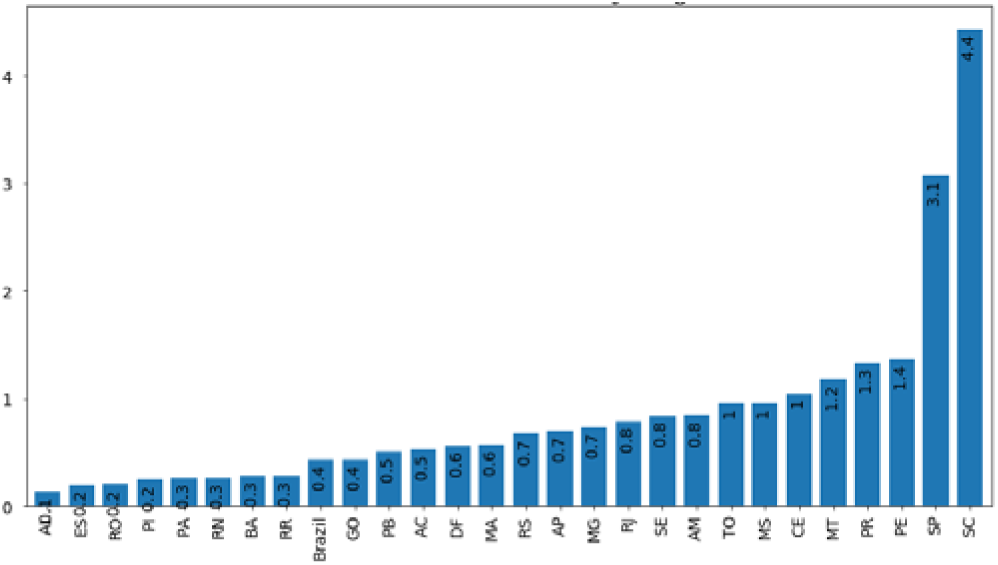
Grotwth rate based on GLM model

**Fig 2.**
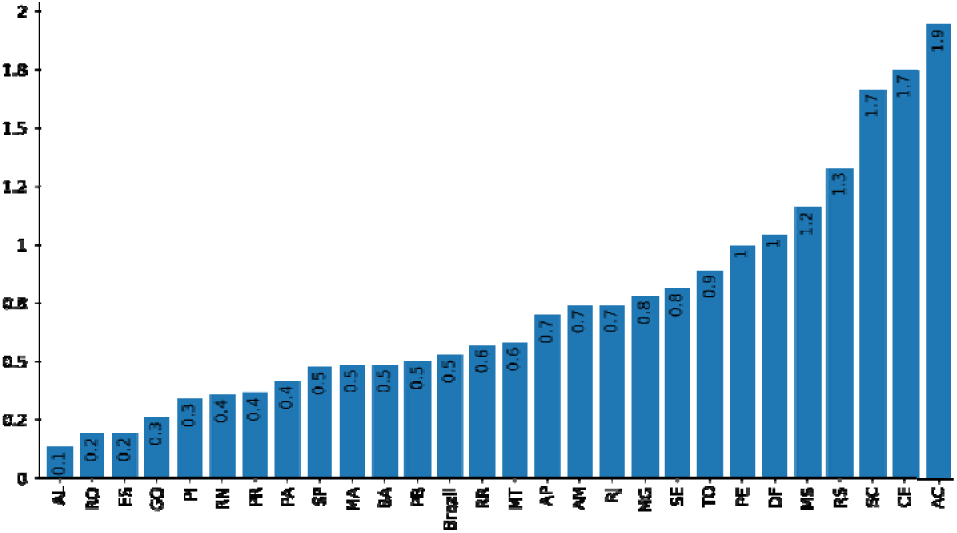
Grotwth rate based on GRM model

**Fig 3.**
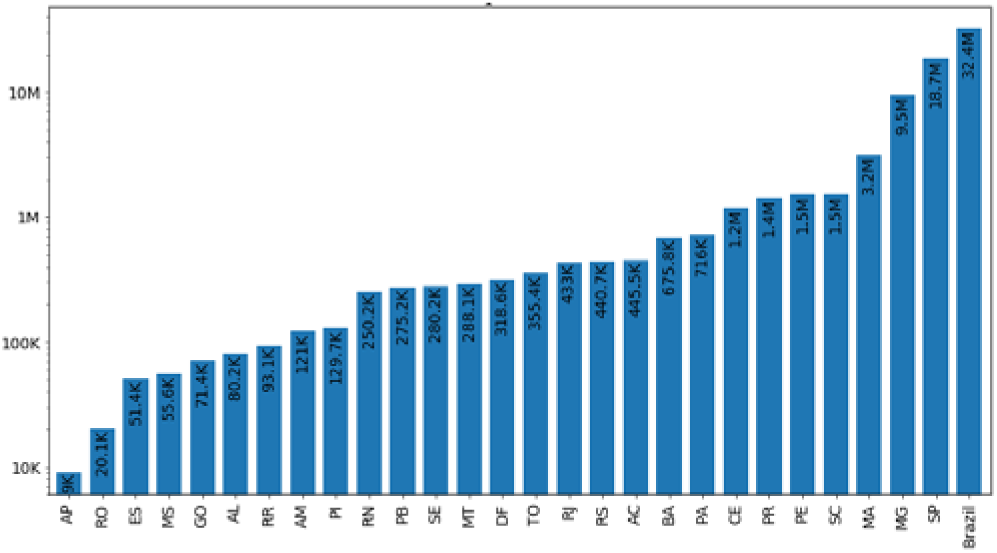
Final epidemic size (K) based on GLM model

**Fig 4.**
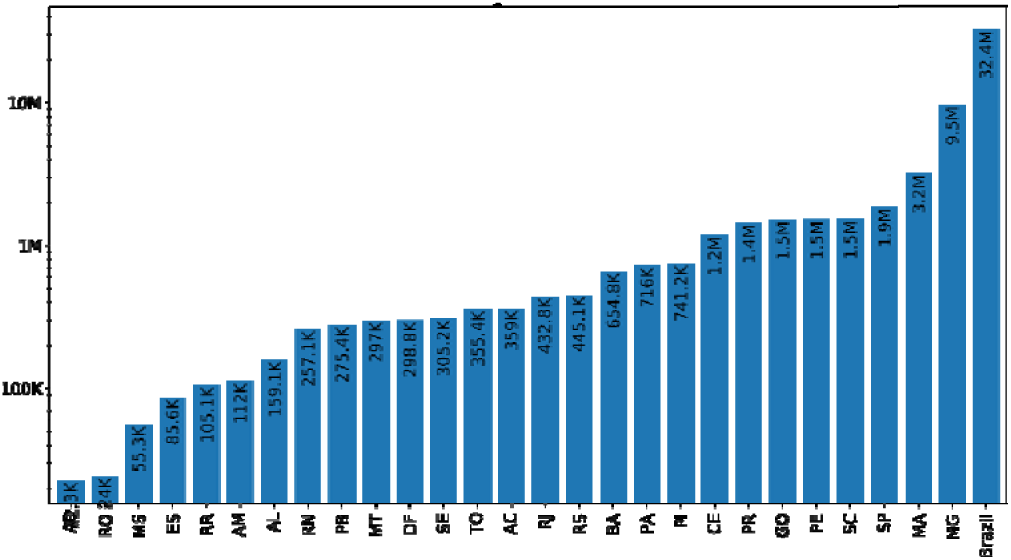
Final epidemic size (K)based on GRM model

### 3.3 Forecast of the total number of cases in 7 days

As stated before, several intervention measures have been adopted by the Federal and State governments. Figures 5–7, below, seek to demonstrate the impact of social mitigation and adoption of new tests, on the trend of increasing cases in Brazil and in the states of Ceará and Pernambuco. After the social mitigation of Brazil, Pernambuco and Ceara, and the top-level health emergency activated in most states on March 25, the transmission has been contained with a relatively fast exponential decay of the growth rate.

**Fig 5.**
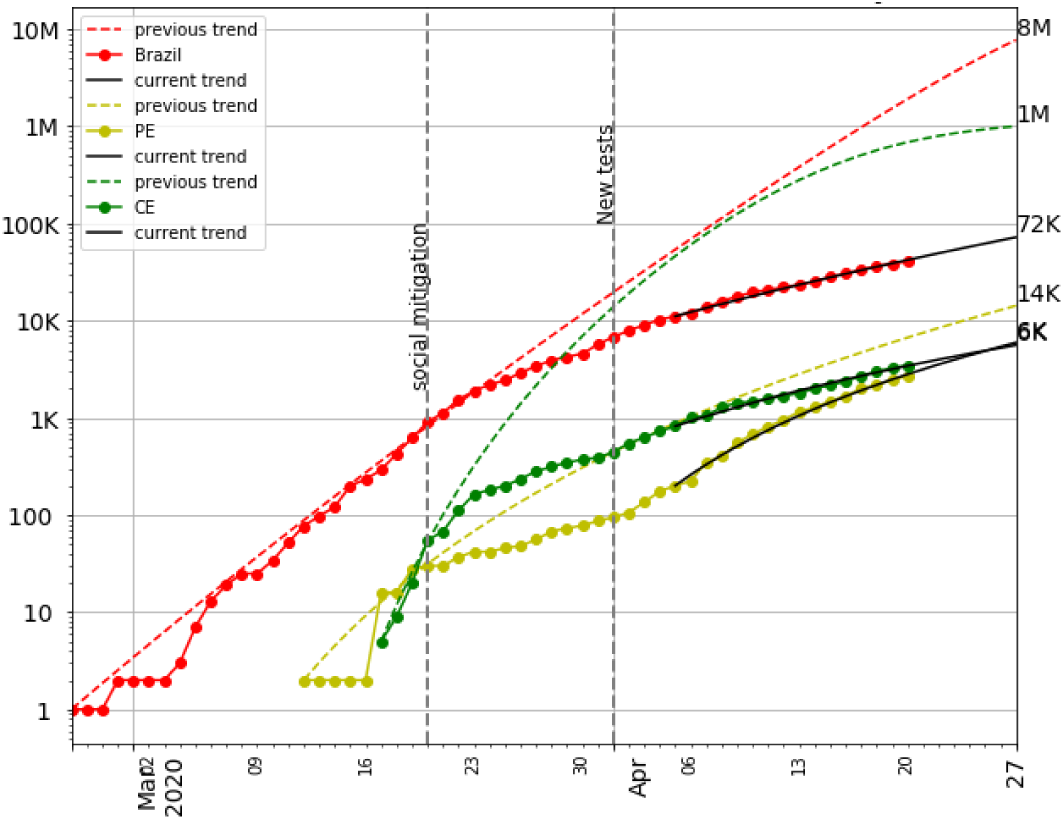
Forecast of the total number of cases in 7 days based on GLM model

**Fig 6.**
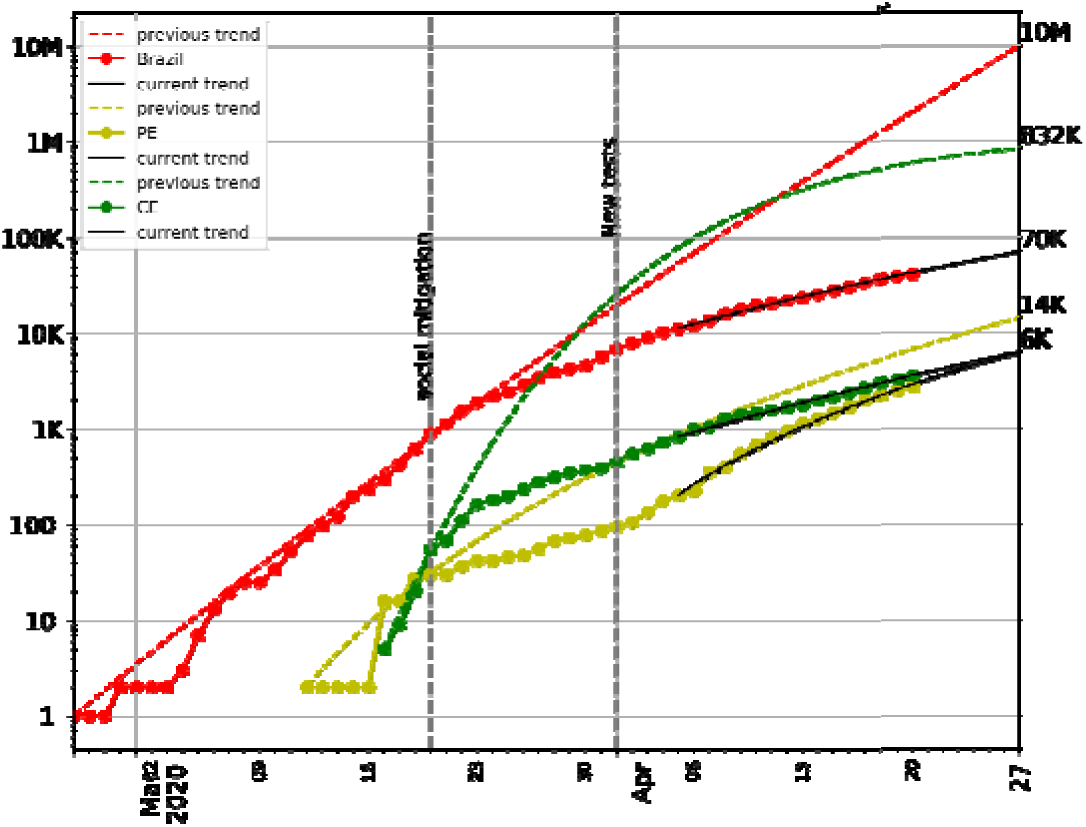
Forecast of the total number of cases in 7 days based GRM model

**Fig 7.**
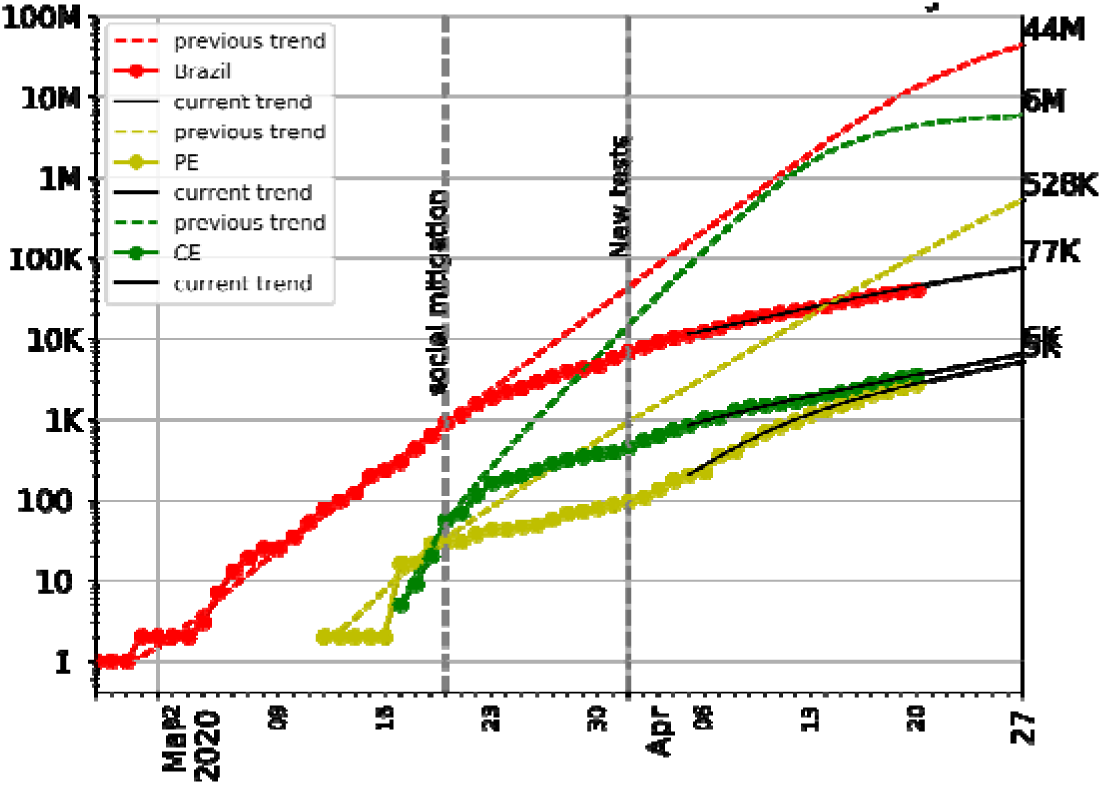
Forecast of the total number of cases in 7 days based SUQC model

We can notice that, firstly there was a convergent analysis between the increasing forecasting of the three models. Secondly, there was a great impact on the growth of cases in the three locations, ranging from 50 to 90%. This is due to the SUQC model, that which presents a greater precision in the estimated number of infected. Nevertheless, this suggests that the epidemic lasts longer in Brazil compared to other states, which may be attributed to intensive control efforts and large-scale social distancing interventions

### 3.4 Short-term predict of the total number of cases in 7 days

Without considering the impact of social mitigation and adoption of new tests, on the trend of increasing cases in Brazil and in the states of Ceará and Pernambuco, we had also conducted a short-term perdiction of the totan numer of cases, in seven days, for all states of Brazil (Figures 8–10). Despite presenting some differences in the estimated values, mainly in the SUQC model, the three models presented the 7 most critical states: Sao Paulo, Rio de Janeiro, Ceara, Pernambuco, Amazonas, Maranhao and Espirito Santo. Among which, to date, April 21, 2020, Amazonas, Ceará and Pernambuco, occupy more than 90% of the ICU beds for Covid-19.

**Fig 8.**
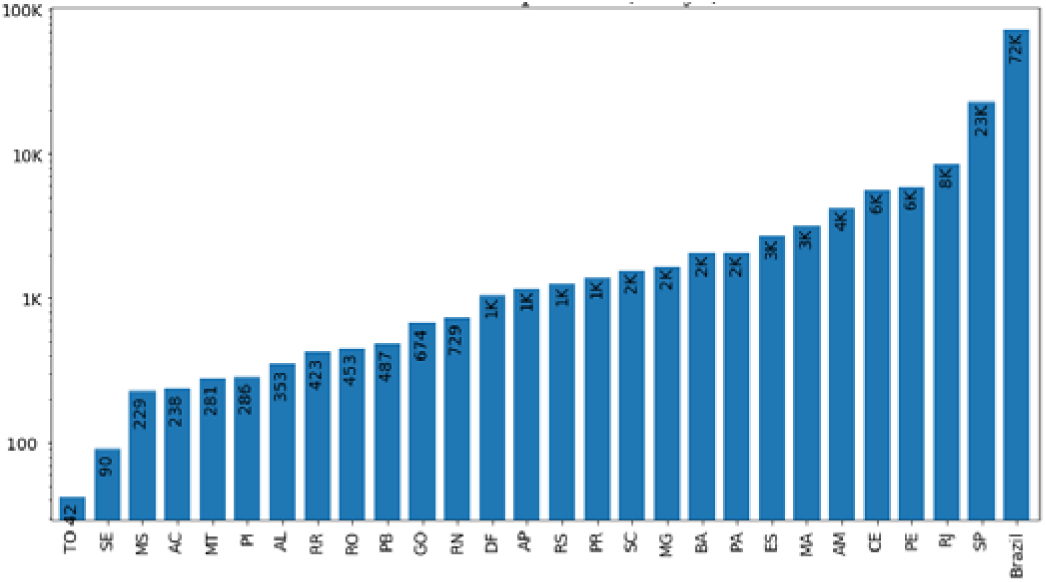
Short-term predict (7 days) based on GLM model

**Fig 9.**
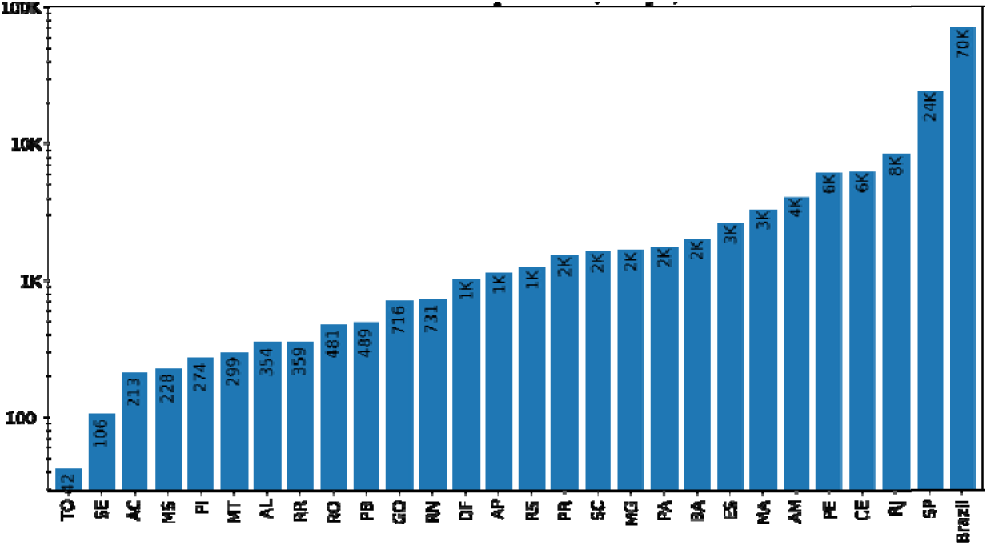
Short-term predict (7 days) based on GRM model

**Fig 10.**
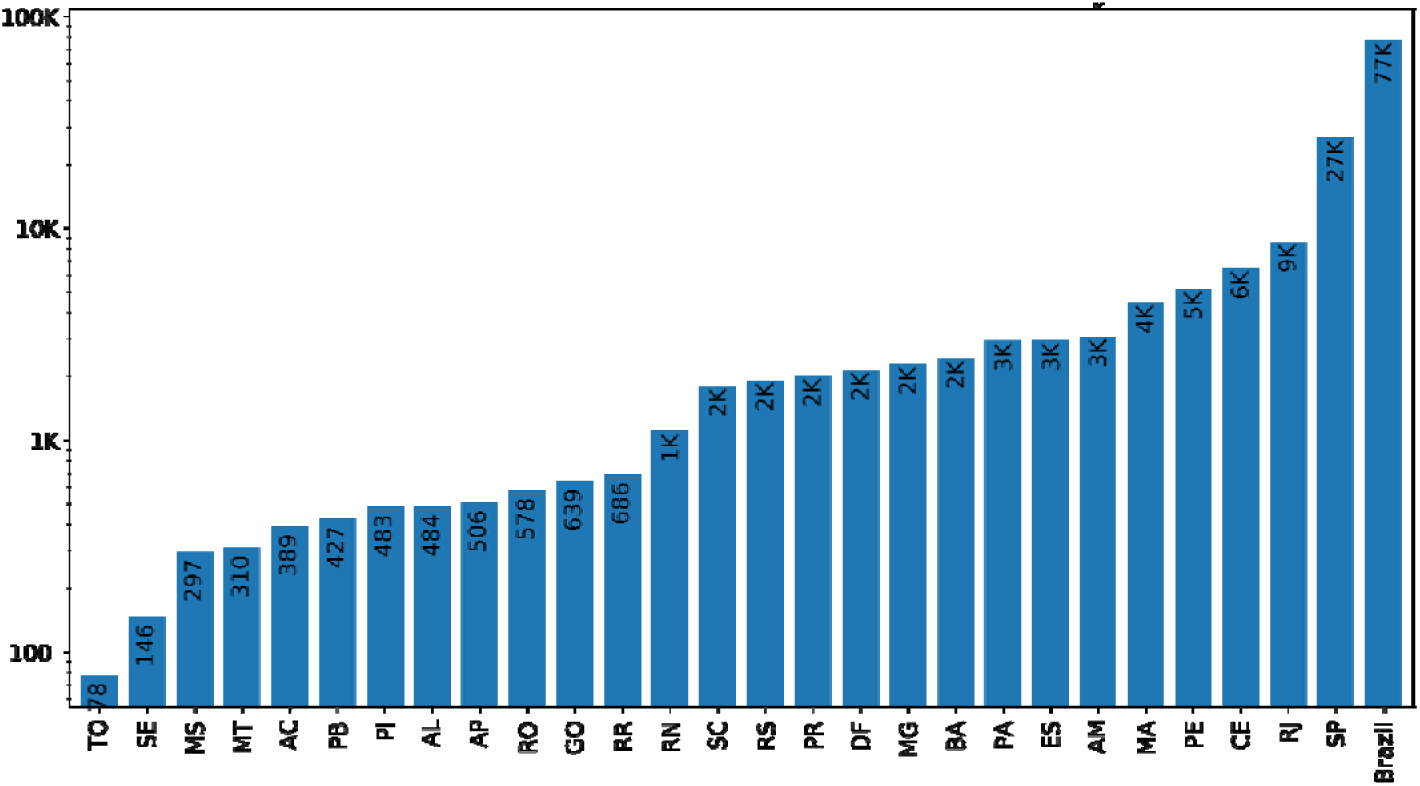
Short-term predict (7 days) based on SUQC model

### 3.5 Interval of days between the beginning and the peak of the epidemic

We had also simulated the infected peak os cases (Figure 11–13). Althgough the result of the three models enphisize greater flattening in the curve, with a manageable number of sick / dead, Pernambuco and Ceara occupy more than 90% of the ICU beds for Covid-19. Besides, according to these results, Ceara calls for more attention.

**Fig 11.**
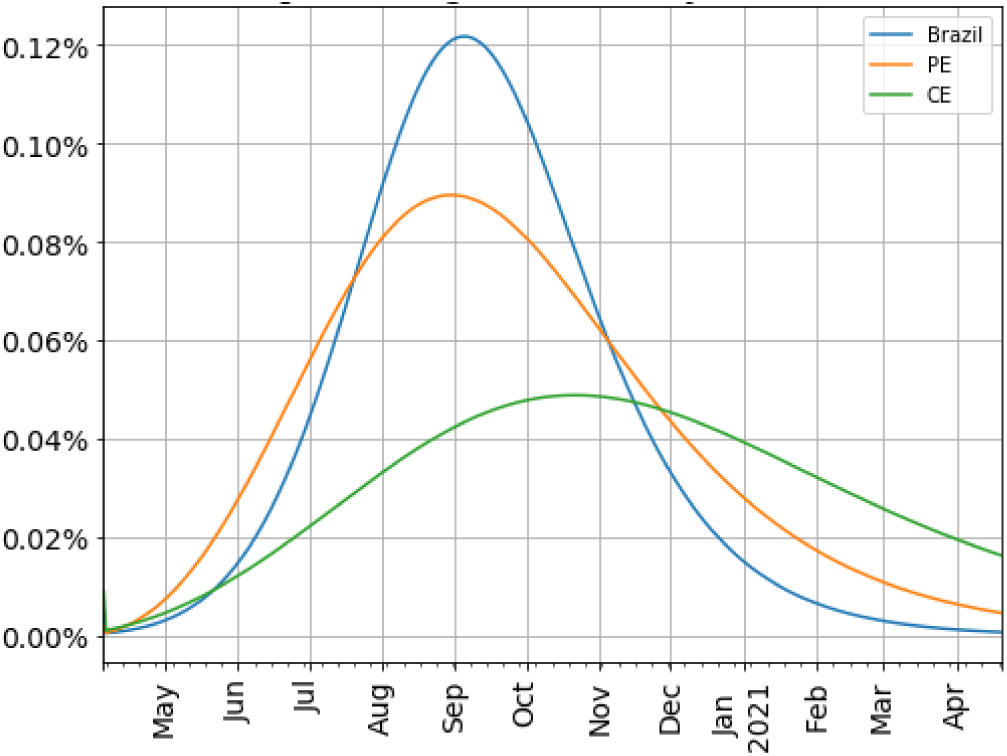
Percentage of daily cases based on GLM model

**Fig 12.**
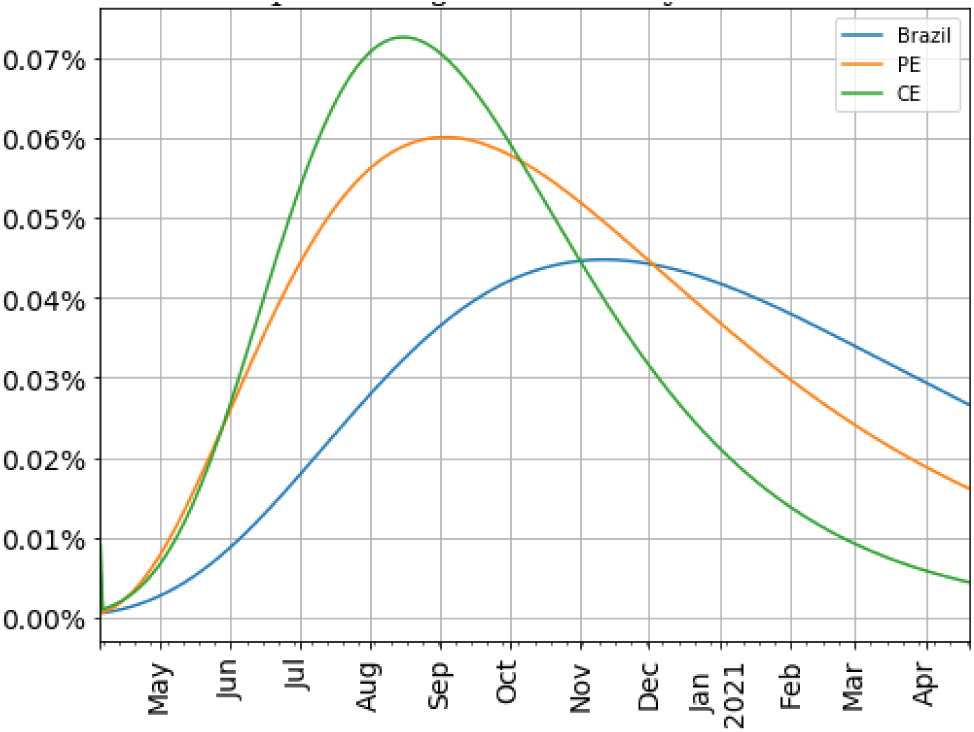
Percentage of daily cases based on GRM model

**Fig 13.**
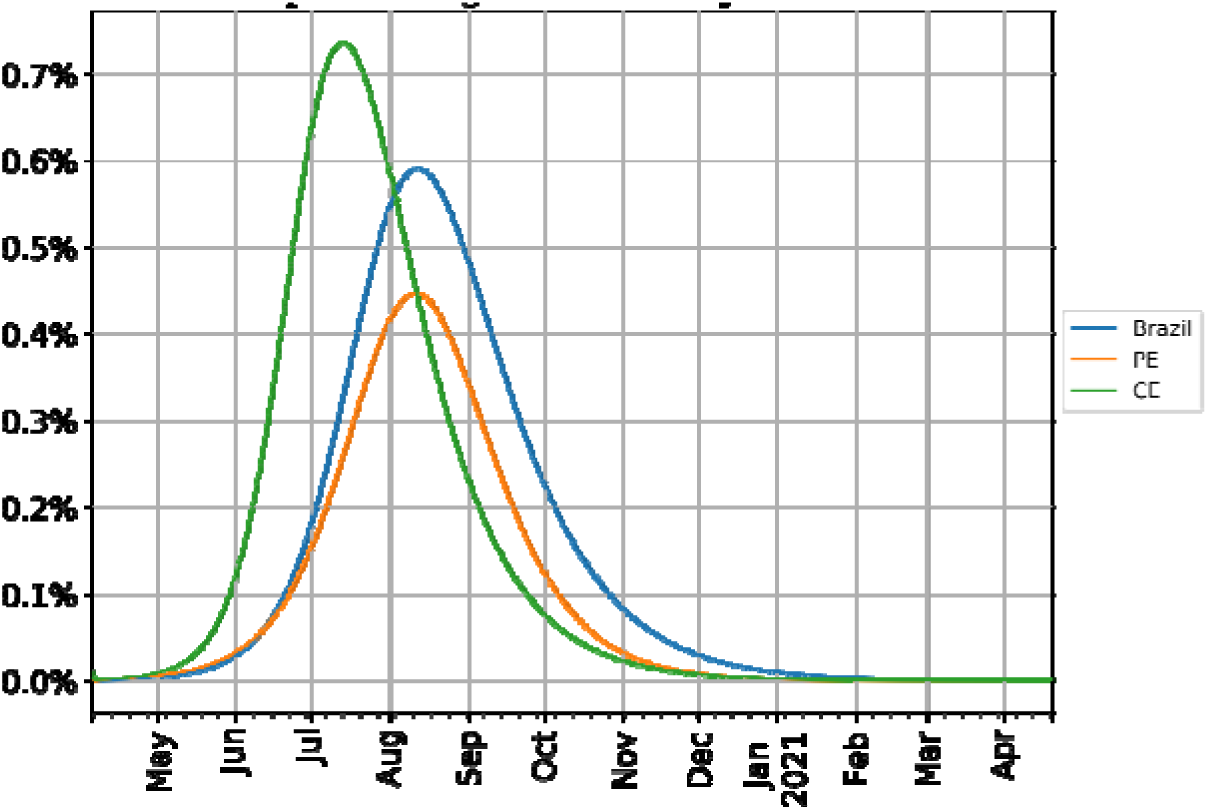
Percentage of daily cases based on SUQC model

### 3.6 Number of days between the beginning and the peak of the epidemic

Take into account all of Brazilian states, we analyzed the number of days between the beginning the start of adjustement and the peak of the epidemic. This information may lead to the understanding that some states are already experiencing the peak of the epidemic, while others are on the verge of that peak.

**Fig 14.**
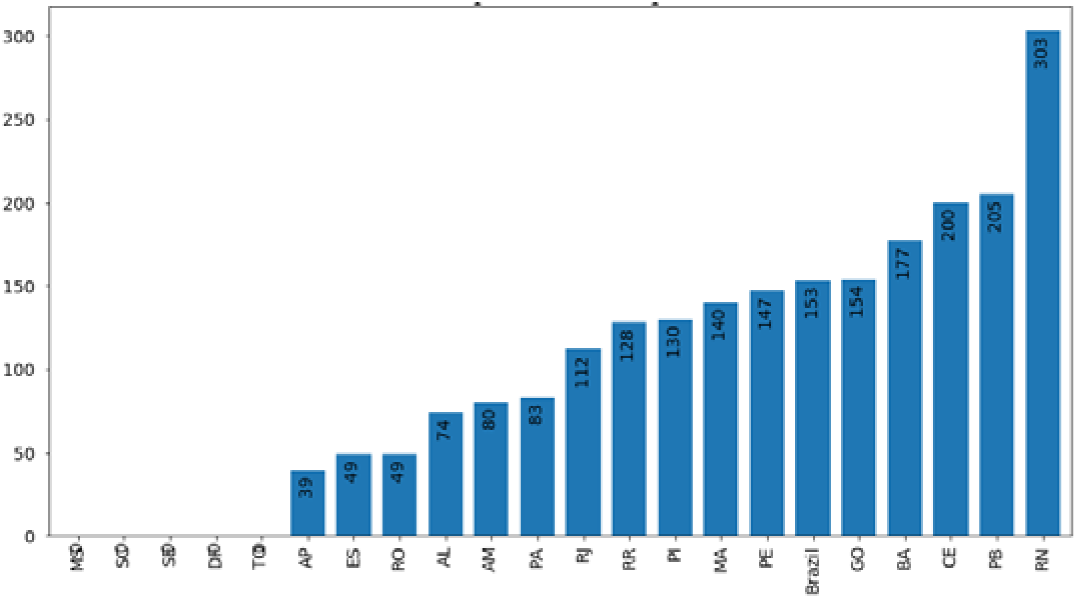
Number of days between start of adjustment and the peak based on GLM model

**Fig 15.**
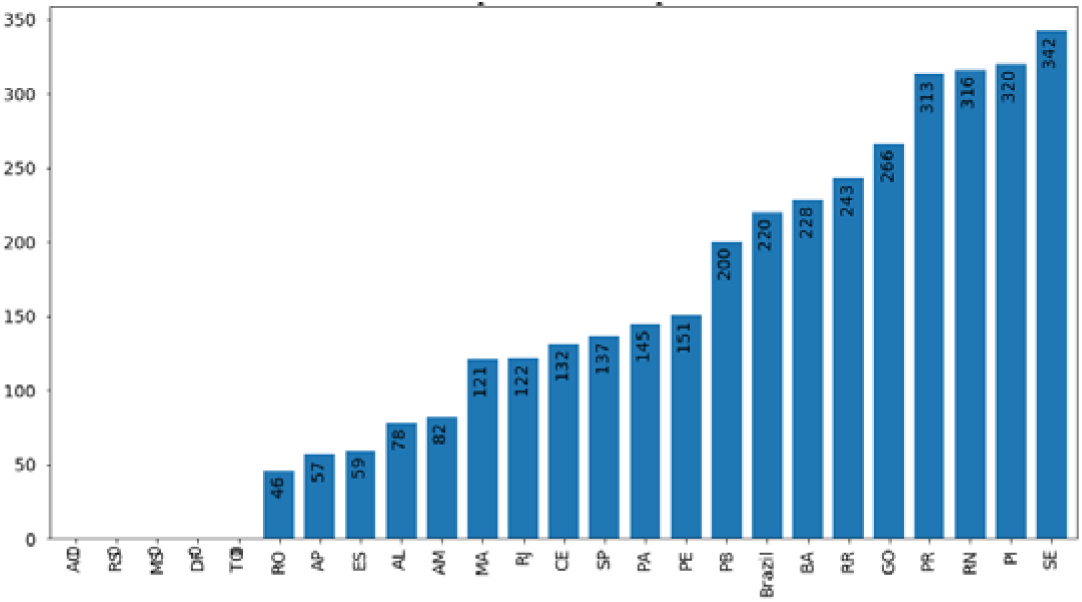
Number of days between start of adjustment and the peak based on GRM model

**Fig 16.**
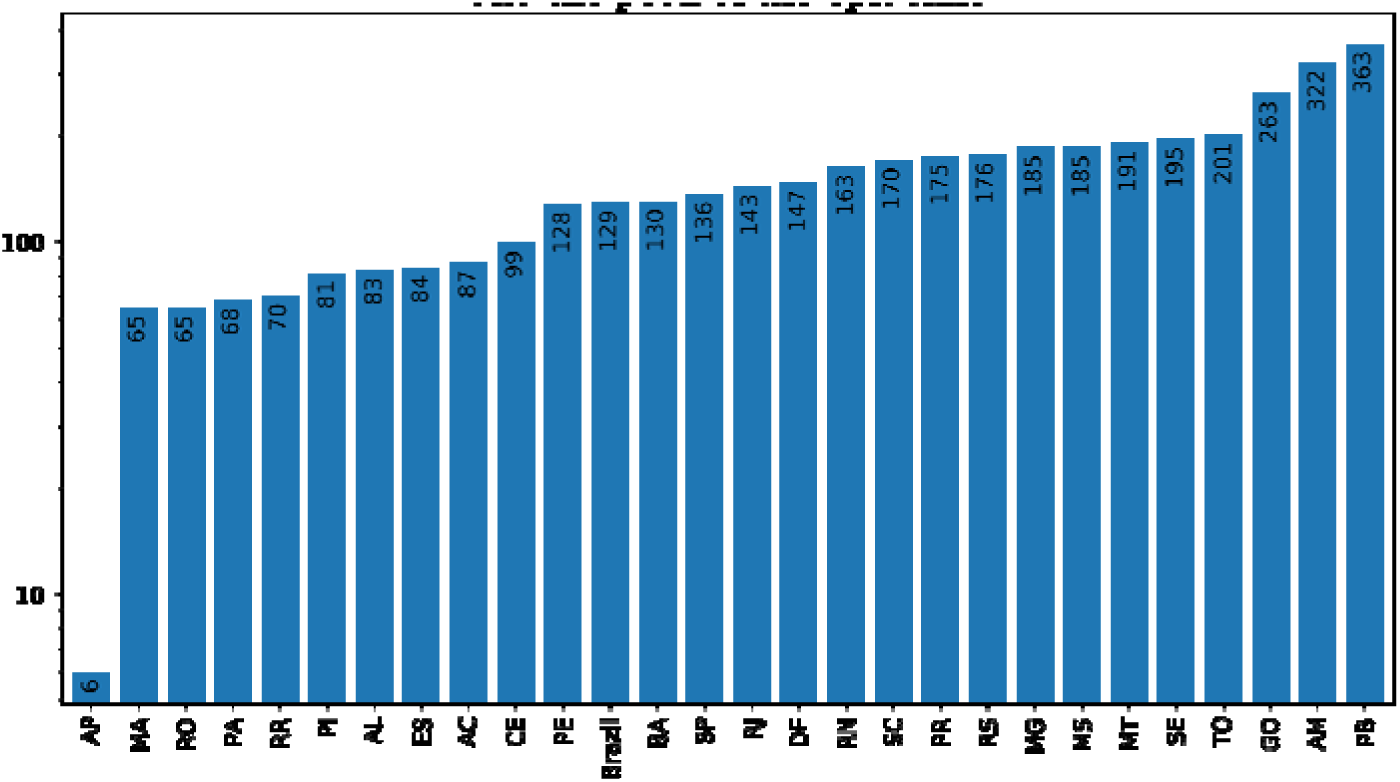
Number of days between start of adjustment and the peak based on SUQC model

### 3.7 Maximum number of cases

Beasides the number of days between the beginning and the peak of the epidemic, we also analyzed the maximum number of daily cases in Brazil and for other states. This information may lead to the understanding that some states are already experiencing the peak of the epidemic, while others are on the verge of that peak.

**Fig 17.**
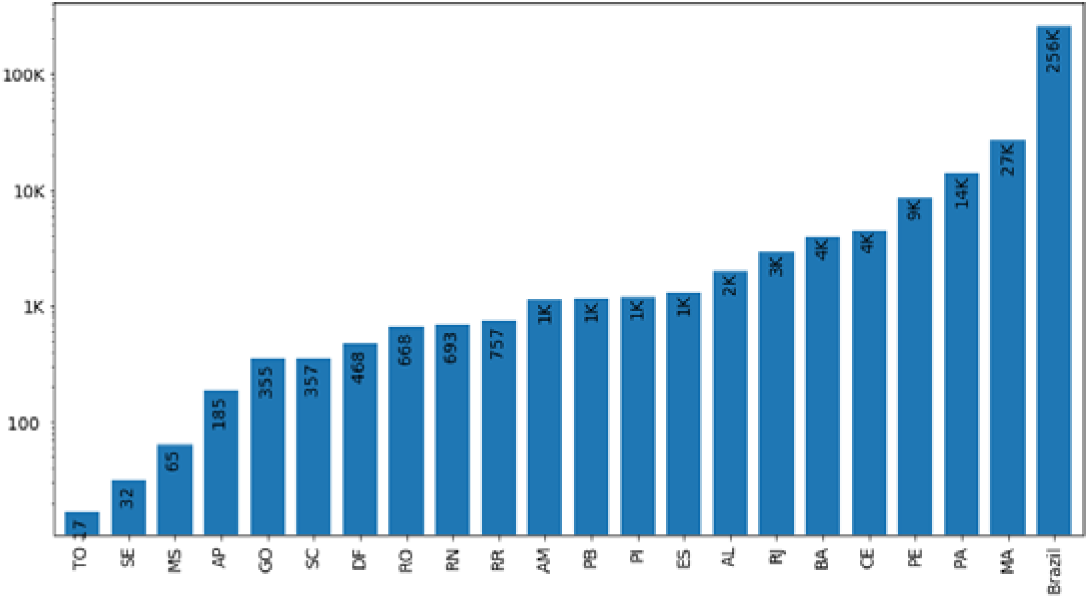
Maximum number of daily cases based on GLM model

**Fig 18.**
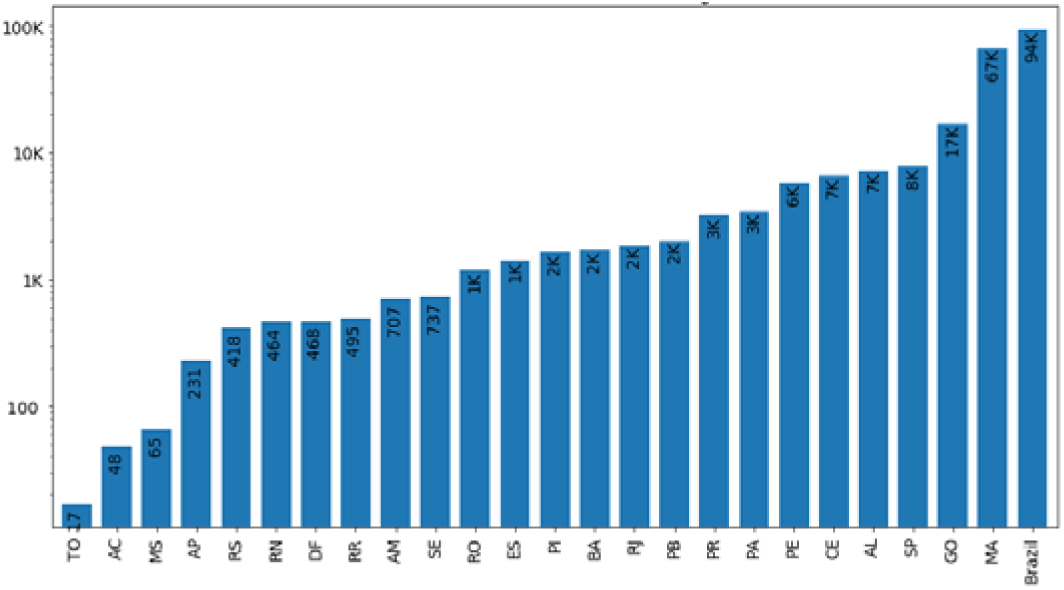
Maximum number of daily cases based on GRM model

**Fig 19.**
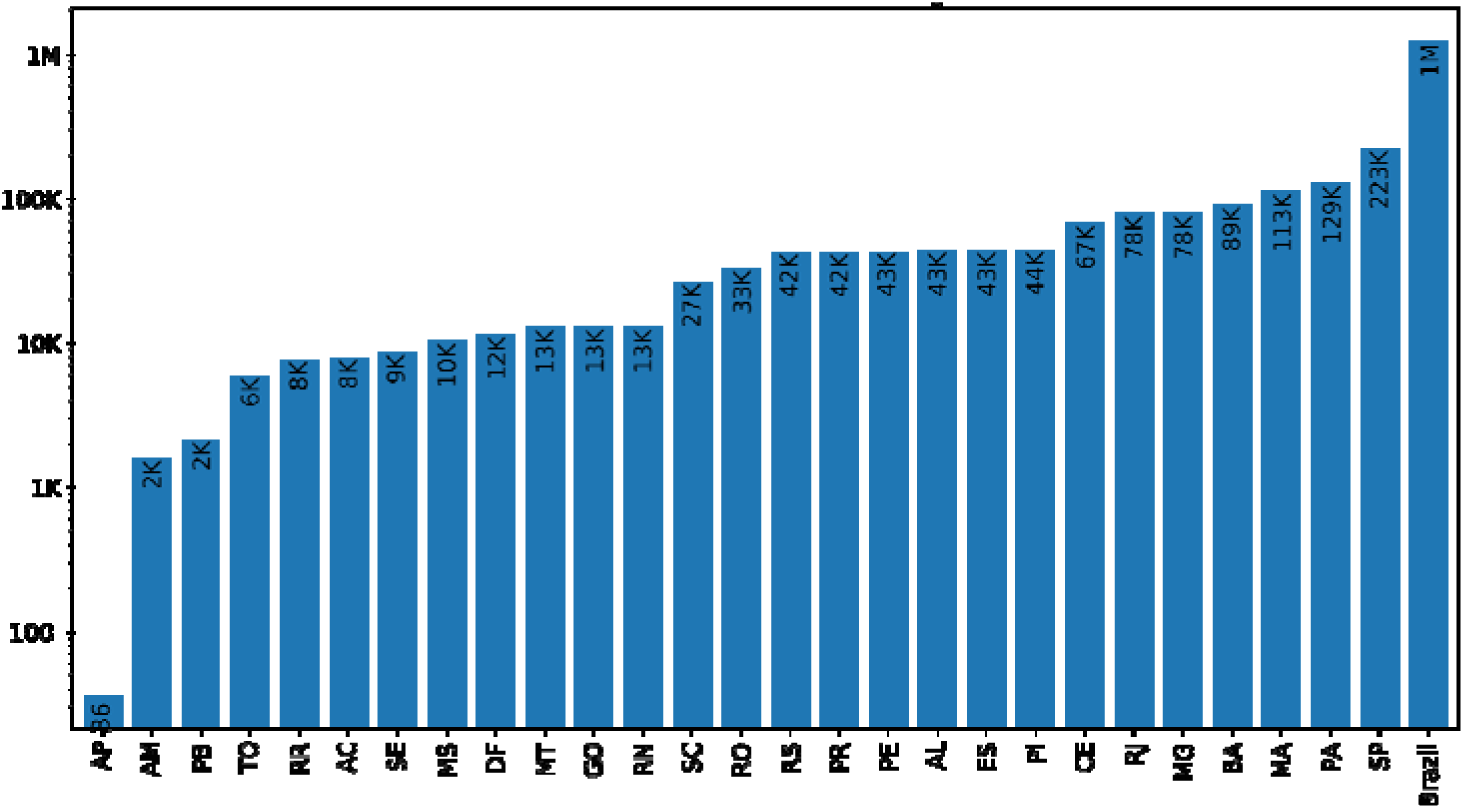
Maximum number of daily cases based on SUQC model

### 3.8 Growth Rate of Reproductive Number

A simulation of the number of reproductions of the virus was made over time, as shown in Figure 20–22, below. It is possible to notice that, in gerenal, Brazil has been showing a downward trend, while the state of PE and CE has a growth trend, mainly after April 13. However, Ceara continues to show greater value for the reprotuvity index in the three models.

**Fig 20.**
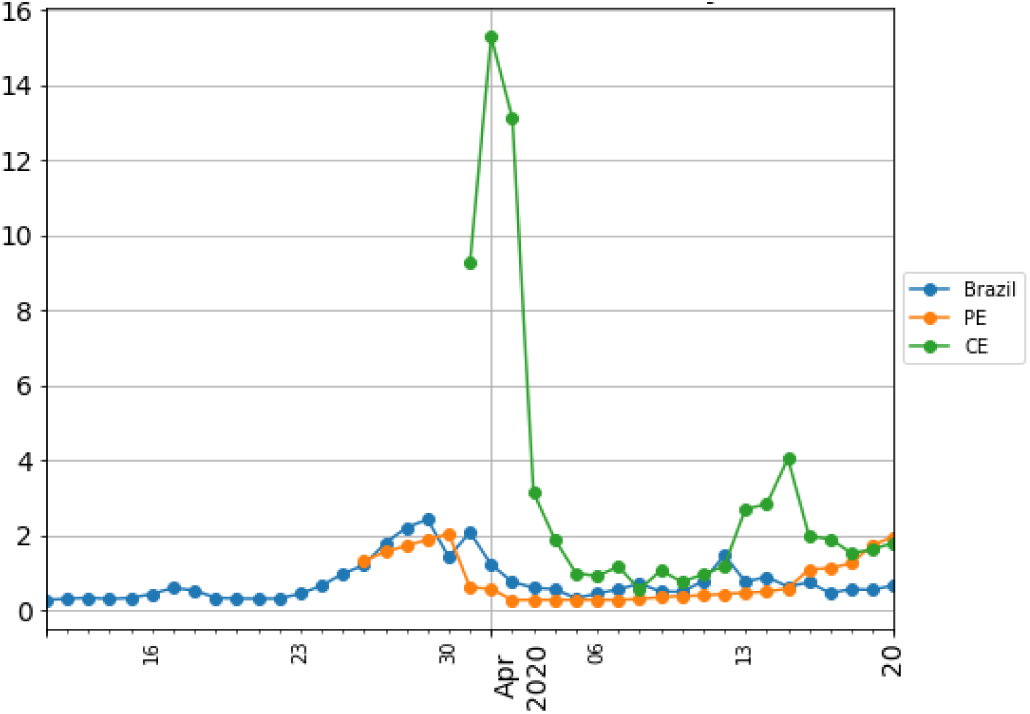
Growth rate (interval of 15 days) based on GLM model

**Fig 21.**
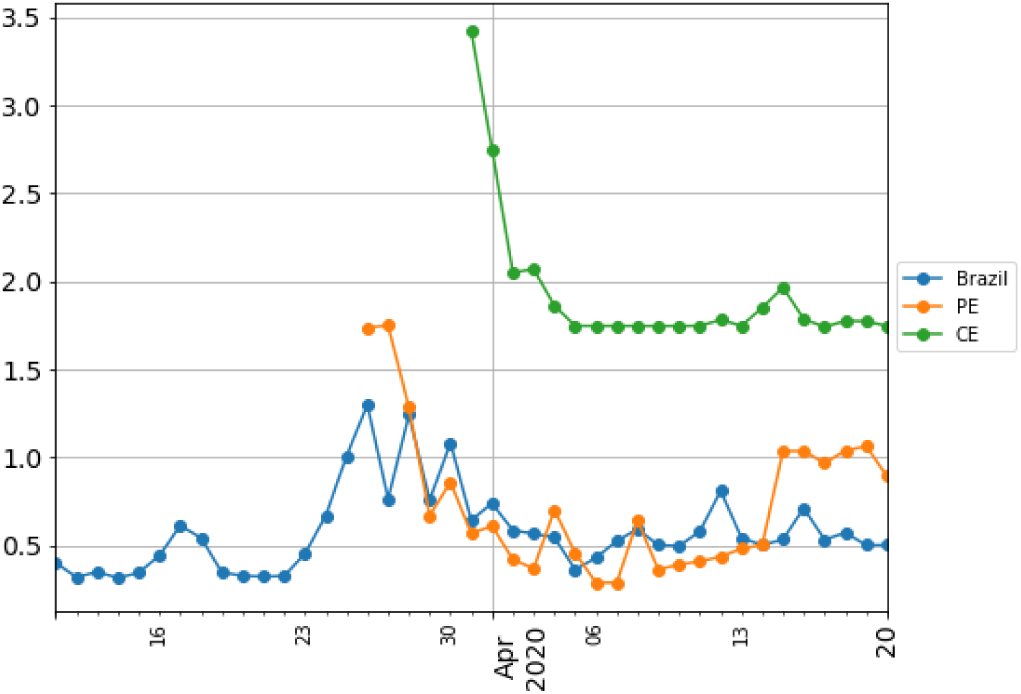
Growth rate (interval of 15 days) based on GRM model

**Fig 22.**
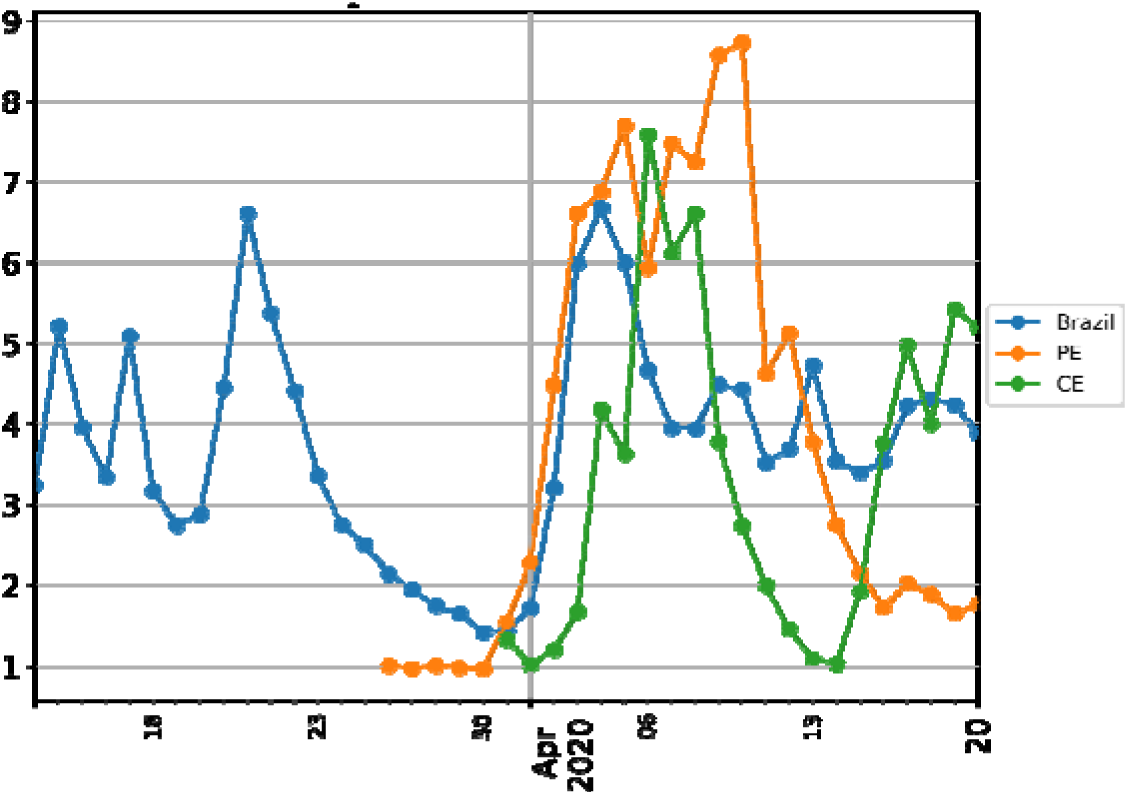
Growth rate (interval of 15 days) based on SUQC model

### 3.9 Total Estimated Death

A simulation of the total number of deaths was made for Brazil and its all states, as shown in Figure 23–25, below, based on the lethality rates. Special attention should be given to Sao Paulo, Pernambuco and Rio de Janeiro states. In fact, until the present date, April 22, according to data from the Ministry of Health, in the ranking of confirmed and fatal cases, Sao Paulo, Rio de Janeiro, Ceara, Pernambuco and Amazonas, lead this list.

**Fig 23.**
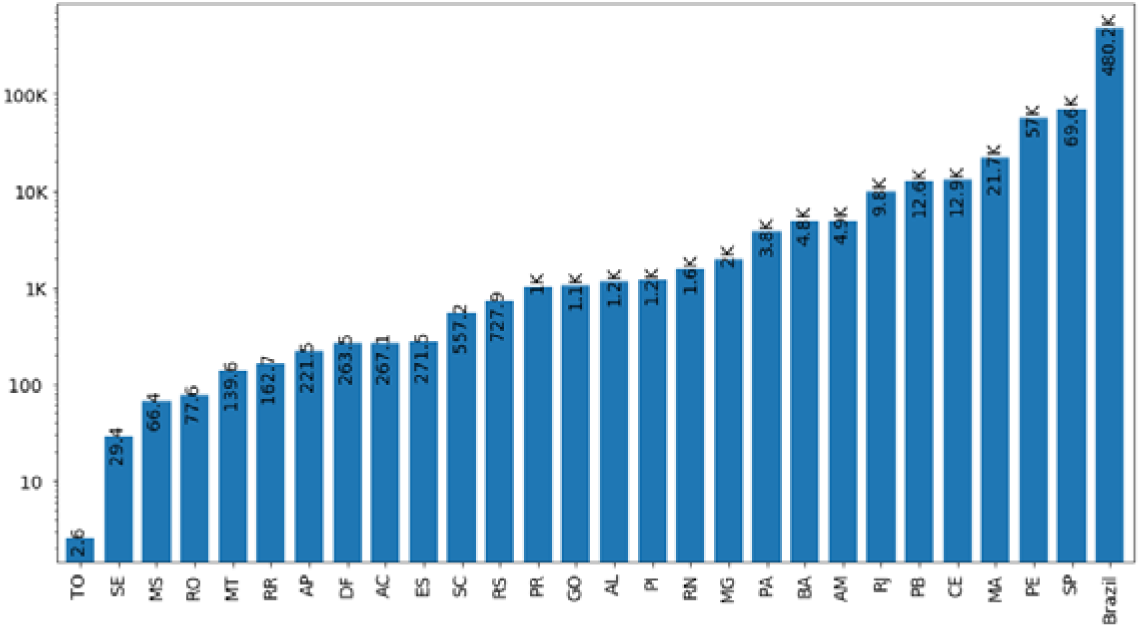
Total estimated deaths based on GLM model

**Fig 24.**
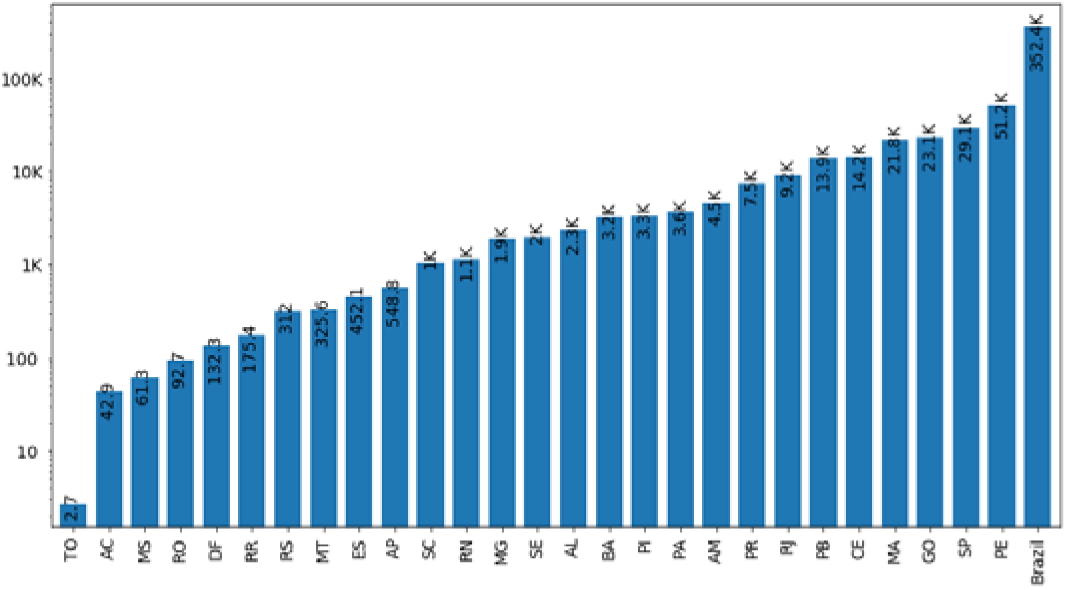
Total estimated deaths based on GRM model

**Fig 25.**
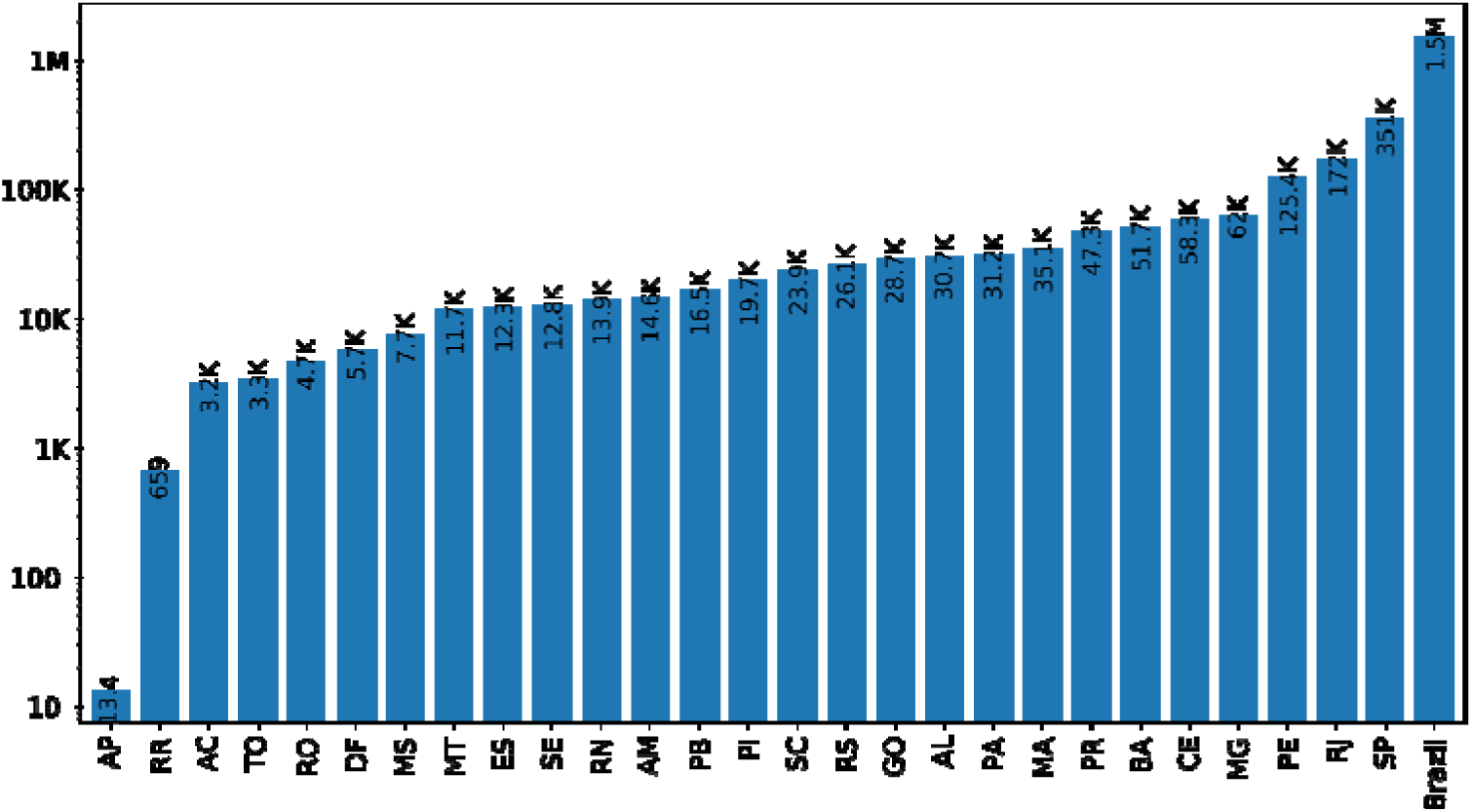
Total estimated deaths based on SUQC model

### 3.10 Final Analysis based on SUQC Model

The SUQC Model, to model COVID-19 cases, is usefull to understand the dynamic of the COVID. Figure 26–28, below, shows the model’s output variables in a 380-day simulation from April 5^th^ for Brazil, Pernambuco and Ceara. As described before, the SUQC model takes into account the number of susceptible people (S), people infected in non-quarantine (U), people infected in quarantine (Q) and the total number of case confirmations (C) from COVID-19, and with them, still the total number of infected people can be estimated (I = U+Q+C).Individual number

**Fig 26.**
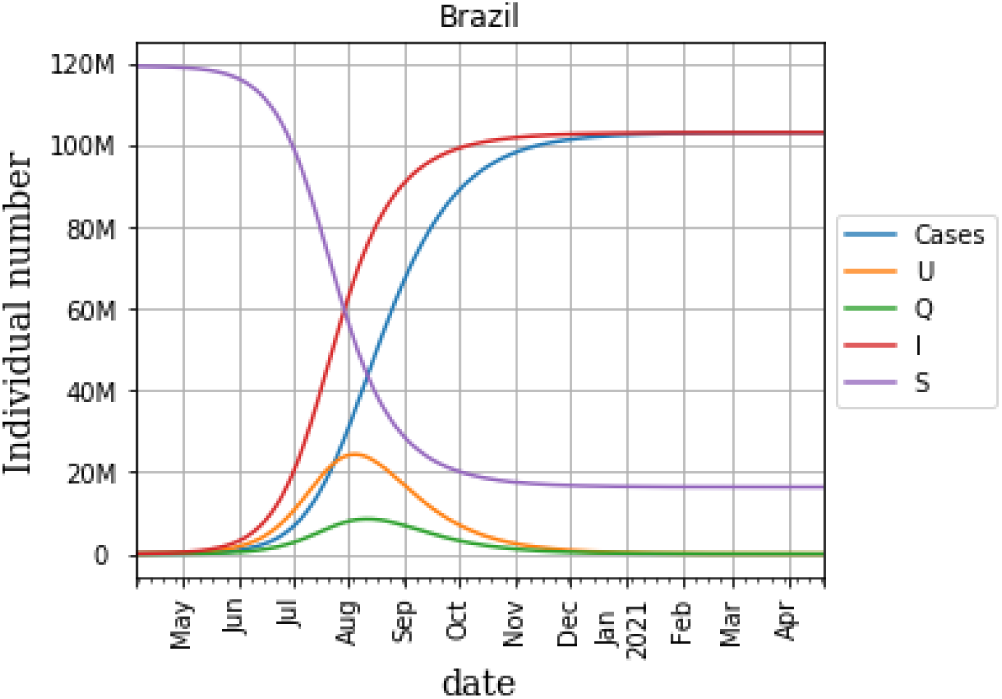
SUQC model of Brazil

**Fig 27.**
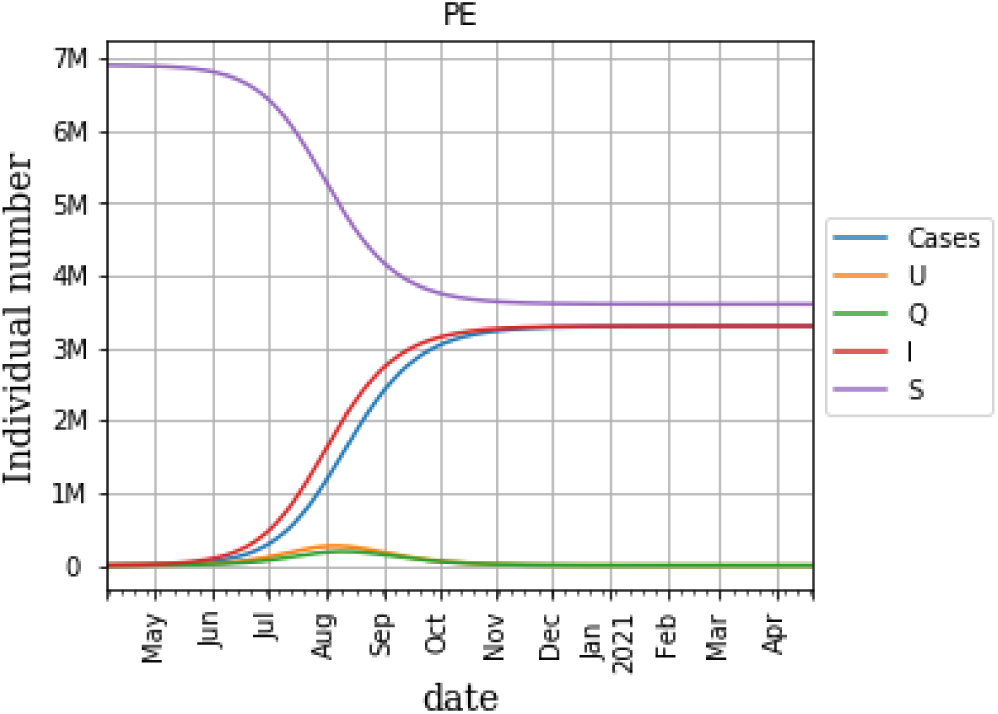
SUQC model of Pernambuco

**Fig 28.**
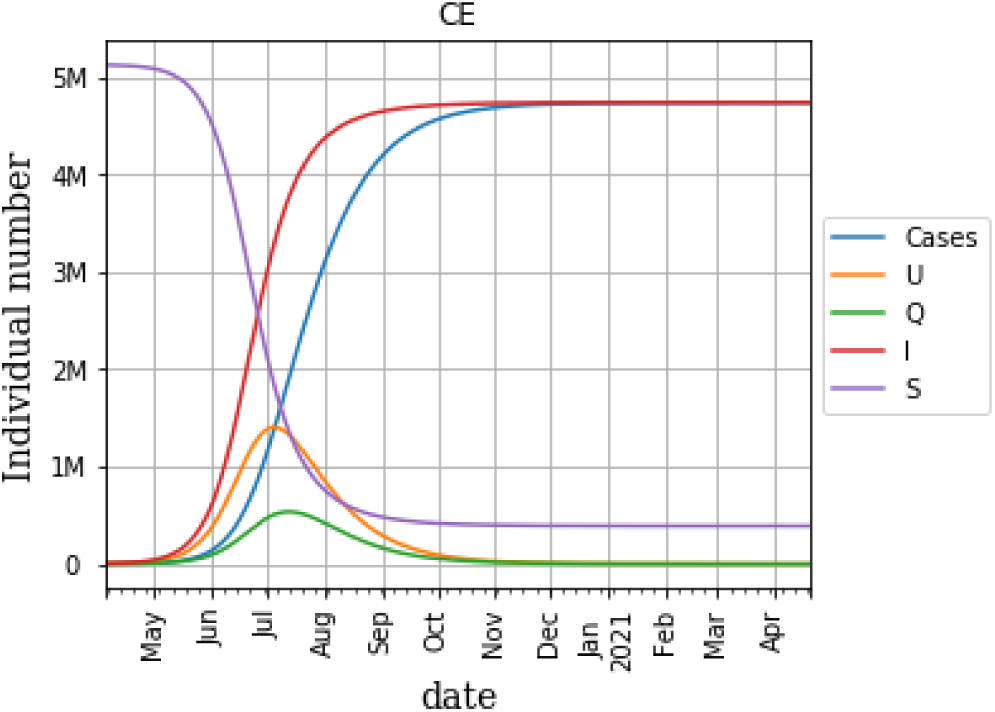
SUQC model of Ceara

As the epidemic runs its course, the susceptible are infected.The Figure 30, below, shows the current lethality of the virus, which was calculated from the ratio between the total number of deaths and the total number of confirmed cases. Among the states reported with the highest lethality rate, Pernambuco, Rio de Janeiro and Sao Paulo. Brazil has a 6.3% lethality rate, that is a rate close to that presented in China (country of origin of the disease).

**Fig 30.**
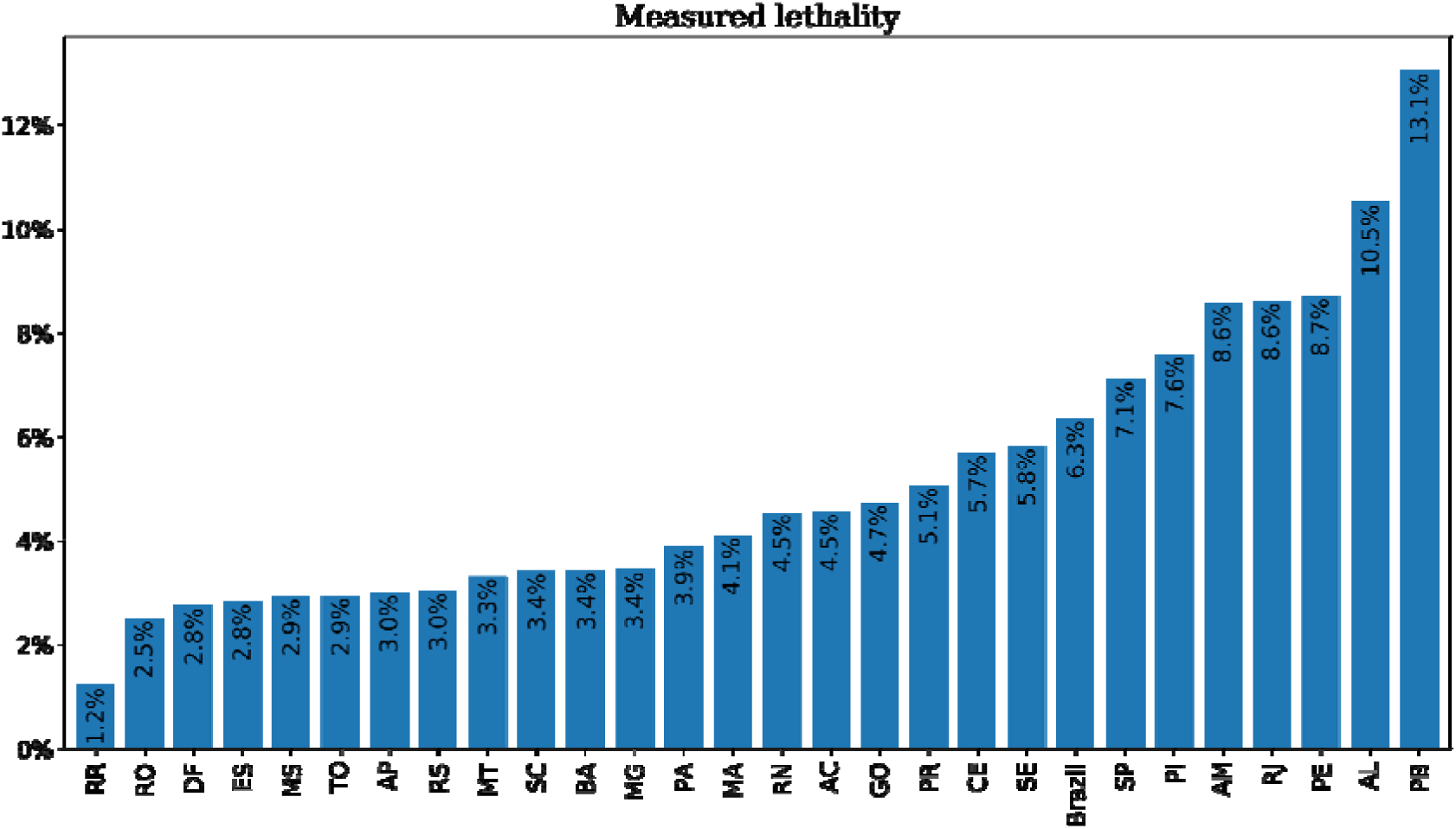
Total deaths per total confirmed cases

By the other side, as the SUQC model is able to provide an estimate of the total number of infected (I), we can estimate the true lethality of the virus, as shown in Figure 31, below:

**Fig 31.**
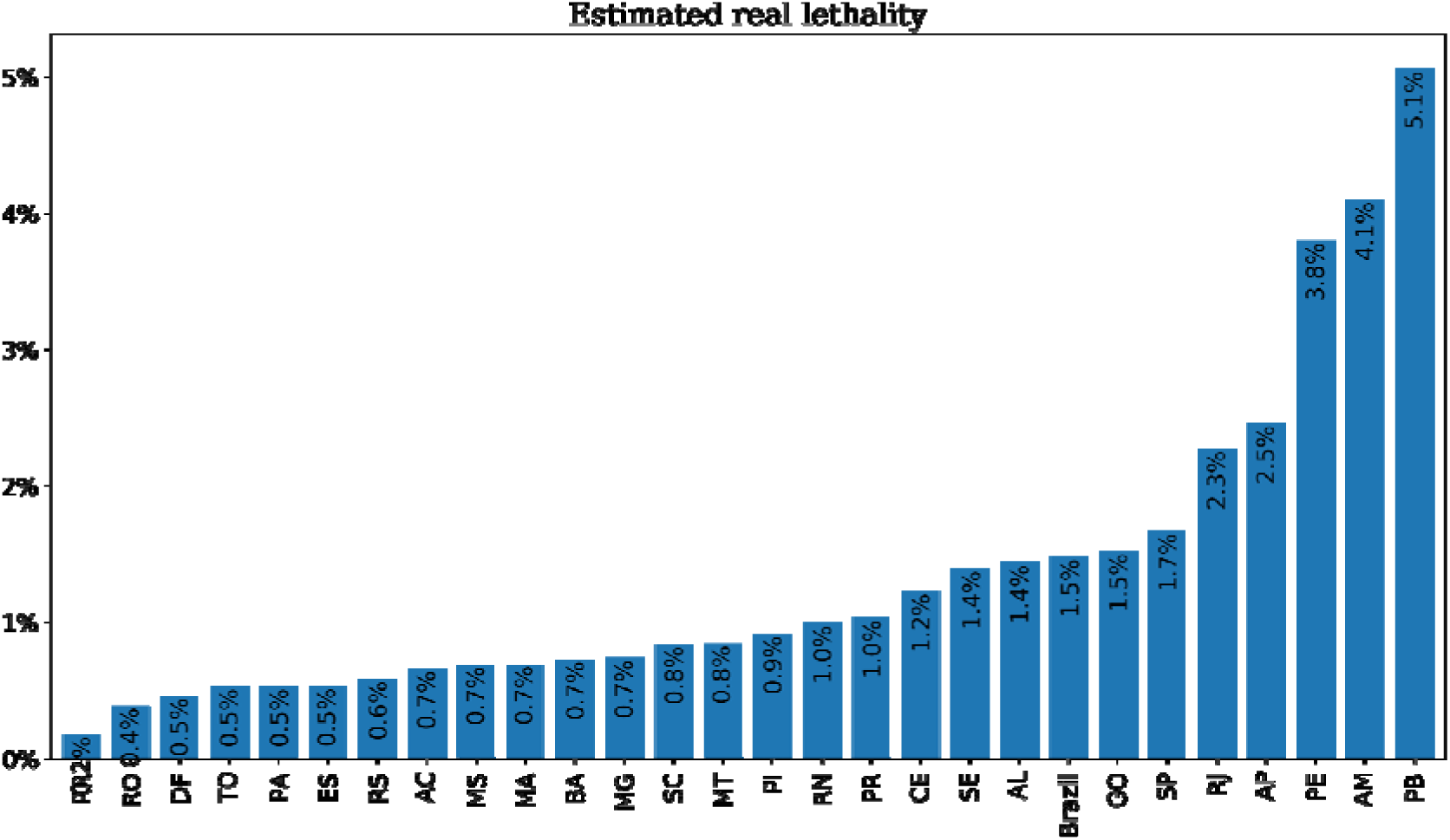
Total deaths per all infected cases

It is noted that, concerning to the lethality calculated from the confirmed cases, there was a reduction of about four times for Brazil. In this new assessment, the country had a 1.5% lethality rate, leaving it very close to the lethality rate of countries with the greatest potential for detecting pre-symptomatic people such as Germany (about 1%). This indicates that the model has a great capacity to estimate the number of pre-symptomatic people who are not in confinement (U). This makes it a crucial tool for making decisions about whether or not to intensify social isolation.

## 4. Discussion and concluding remarks

In this paper, we provide timely short-term forecasts of the cumulative number of reported cases of the 2019-nCoV epidemic in Brazil and the state of Pernambuco and Ceara, northeast Brazil, based on the GLM, Richards and SUQC model. By using this procedure, we refit our proposed dynamics transmission model to the data available until April 16th, 2020 and re-estimated the daily reproduction number, which implies the evolving epidemic trend. Overall, our findings supports several other studies (J. Zhang et al., 2020), in which indicate that intervention measures might contribute to interrupt local transmission.

The forecasts presented are based on the assumption that current mitigation efforts will continue.

## Data Availability

Data from https://saude.gov.br

## Acknowledgments

This research is partially supported by FADE and UFPE. The authors are also grateful to Pernambuco Government for providing data which made the research possible.

